# Elevated antibody titers in Abdala vaccinees evaluated by Elecsys® anti-SARS-CoV-2 S highly correlate with UMELISA SARS-CoV-2 ANTI RBD, ACE-2 binding inhibition and viral neutralization assays

**DOI:** 10.1101/2021.10.18.21265169

**Authors:** Gilda Lemos-Pérez, Sheila Chávez-Valdés, Hany González-Formental, Giselle Freyre-Corrales, Amalia Vázquez-Arteaga, Beatriz Álvarez-Acevedo, Lismary Ávila-Díaz, Ricardo U. Martínez-Rosales, Yahima Chacón-Quintero, Edelgis Coizeau-Rodríguez, Ariel Palenzuela-Díaz, Enrique Noa-Romero, Gerardo E. Guillén-Nieto

**Author notes:** **Corresponding author**: Gilda Lemos Pérez MSc., Biomedical Research Direction, Center for Genetic Engineering and Biotechnology, 27 Ave. 31 E/ 158 y 190, La Habana 10600, Cuba.

## Abstract

SARS-CoV-2, a recently emerged coronavirus, is causing high morbidity and mortality worldwide since December 2019, posing an enormous health, social and economic problem. Obtaining effective treatments that can diminish deaths and sequelae and vaccines to slow or prevent viral transmission, and reduce disease severity and/or death are of utmost importance. Abdala is a Cuban vaccine based on the recombinant RBD subunit of the spike protein expressed in *Pichia pastoris* yeast. It demonstrated high efficacy (92.28 %) in phase III clinical trials for reducing transmission, and more than 90% effectiveness in reducing disease severity and mortality. Antibody titers were evaluated in 42 Abdala vaccinees using the Elecsys® Anti-SARS-CoV-2 S test. Fifteen days after immunization, sera from vaccinees showed high antibody titers (median of 1595 U/mL). The results obtained in this study also demonstrate correlation between the Cuban test UMELISA SARS-CoV-2 ANTI RBD used during the clinical trials and Elecsys® test results.

**Highlights:**

- Fifteen days after immunization with the Cuban Abdala vaccine, sera from vaccinees showed high antibody titers (median of 1595 U/mL).
- There was high correlation between the Cuban test UMELISA SARS-CoV-2 ANTI RBD used during the vaccine clinical trials and Roche’s Elecsys® test results.
- Abdala vaccinees reached antibody titers by the Elecsys® Anti-SARS-CoV-2 S test comparable to those of Pfizer/BionTech vaccine using the same test.

## Introduction

COVID-19 is a severe disease produced by a new coronavirus (SARS-CoV-2), whose first victims were reported in Wuhan, China, in late 2019. This disease rapidly spread to other provinces in China and to the rest of the world [1]. By the end of January 2020, The World Health Organization (WHO) declared this disease a global health emergency [2], and it was recognized as a pandemic in March 2020 [3]. On March 23, confirmed SARS-CoV-2 cases exceeded 3,000 globally; the disease was present in 185 territories in six continents. The death total was 14,502. Together with the well-known containment measures for respiratory infection control, testing of suspected cases and contacts (with or without symptoms) by rRT-PCR was highly recommended [4]. On March 27, 2020, the USA became the country with the most confirmed cases, surpassing China. A few days later (on April 3), the number of confirmed cases globally exceeded one million and global deaths surpassed 56,000 [5]. In Cuba, the first COVID-19 case was confirmed on March 11 [6].

The rapidly growing infection rate of SARS-CoV-2 worldwide that poses enormous health, social and economic problems, triggered a race for developing diagnostics, therapeutic agents and vaccines, which might ultimately slow or prevent viral transmission. The entire genome structure of SARS-CoV-2 (COVID-19 etiologic agent) was made available since early January 2020 [7]. It is almost identical to SARS-CoV except for some additional protein genes and its host receptor, the ACE-2 (angiotensin-converting enzyme 2) protein that allows entry into the cell. Among the structural proteins of SARS-CoV-2, the spike (S) and nucleocapsid are the main immunogens used in diagnosis. The S protein contains two subunits, S1 and S2. The S1 subunit contains a receptor binding domain (RBD), which is responsible for recognition and binding to the cell surface receptor, making it the main target for vaccine development. The S2 subunit contains other basic elements necessary for membrane fusion [8,9].

Several approaches have been used for COVID-19 vaccine development: the classic approach of inactivated (Sinopharm (BBIBP-CorV) and Sinovac, CoronaVac Chinese vaccines) and virus vector vaccines, such as the Oxford/AstraZeneca AZD1222 (UK), Janssen (Johnson & Johnson (Ad26.COV2.S, USA) and Gamaleya’s Sputnik V (Gam-COVID-Vac, Russia). The newest are the mRNA vaccines Moderna (mRNA-1273, USA) and Pfizer/BioNTech (BNT162b2, Germany), approved for human use for the first time. Protein subunit vaccines like Novavax (NVX-CoV2373, UK) have also entered the market [10].

Cuba worked on five vaccine candidates [11]. The Cuban Center for Genetic Engineering and Biotechnology (CIGB) obtained Abdala®, a Cuban vaccine also named CIGB-66, based on the recombinant RBD subunit of the spike protein produced in *Pichia pastoris* yeast [12]. Its first clinical trial, a Phase I/II trial, began on December 7, 2020 [13] and the Phase III trial was started on March 22, 2021 [13] with a 92.28% efficacy result [15, 16]. At the same time, since the city of Havana became the epicenter of community transmission in Cuba, a vaccine intervention with Abdala was carried out in this city from March 29 to September 30, 2021 [17]. The vaccine was approved for emergency use on July 9 [18,19]. Recent reports have shown that the Abdala vaccine was more than 90% effective against severity and death, despite the prevalence of the Delta variant of SARS-CoV-2 [20].

There are currently two ongoing clinical trials, one in pediatric ages 3 to 18 years and another in convalescents, in whom the immune response is boosted with a single dose of the Abdala or the nasal vaccine candidate Mambisa (CIGB-669, Cuba), one of the nine on trial using this route of administration [21]. The latter completed a successful Phase I trial in healthy and convalescent subjects [22, 23].

High antibody titers, primarily IgG, correlate with good virus neutralizing levels and thus protective immunity [24,25].

The objective of this study was to evaluate serum samples from Abdala vaccinees using the Elecsys® Anti-SARS-CoV-2 S system (Roche Diagnostics, Switzerland) and correlating the results with those previously obtained with the Cuban test (UMELISA SARS-CoV-2 ANTI RBD).

## Materials and methods

### Vaccine

The Cuban Abdala vaccine (CIGB-66) developed by the CIGB (Center for Genetic Engineering and Biotechnology, Cuba) was administered as immunogen [12,19,20].

### Serum samples

The study was carried out using sera collected from 42 individuals immunized with the Abdala vaccine candidate. In total 126 sera were used: prior to immunization (Pre), and fifteen days post both the second and the third (last) immunizations (Post). Collected sera were stored at -20°C until evaluation.

### Ethics

Subjects gave their consent after being informed that the immediate use of the samples was to determine post vaccination antibody titers and there were possibilities of their use in other related research. Confidentiality of the donors’ personal data was ensured. The study was approved by CIGB Ethics Committee in Havana, Cuba.

### SARS-CoV-2 total antibody quantifications

#### Elecsys® Anti-SARS-CoV-2 S assay

SARS-CoV-2 antibodies against the Spike RBD viral protein were determined using the quantitative Elecsys® Anti-SARS-CoV-2 S test on the Cobas e411 Analyzer (Roche Diagnostics). The test is a double-antigen sandwich electro-chemiluminescence immunoassay, which uses streptavidin-coated microparticles to separate bound from unbound substances prior to applying voltage to the electrode. This assay has a measuring range of 0.40-250 U/mL (up to 2500 U/mL with on-board 1:10 dilution), with a concentration ≥0.80 U/mL considered positive. The manufacturer specific U/mL of the Elecsys® Anti-SARS-CoV-2 S assay can be considered equivalent to the BAU/mL of the first WHO International Standard for anti-SARS-CoV-2 immunoglobulin [26].

#### UMELISA SARS-CoV-2 ANTI RBD assay

Class IgG antibodies were quantified by UMELISA SARS-CoV-2 ANTI RBD, approved by CECMED (Cuban Center for State Control of Medicines and Medical Devices) and commercialized by the ImmunoAssay Center (Cuba) [27]. The test is an automated fluorescence-based quantitative assay that uses ultramicro-ELISA plates coated with the RBD fragment of the virus S protein as solid phase. Briefly, 10 µL of 1:100 pre-diluted samples are incubated in the wells of the strips for 30 minutes. After a wash step, biotin-bound anti-human IgG monoclonal antibody is then added, which will bind to the antibodies fixed in the previous step. A further wash removes the unreacted biotinylated antibodies. A streptavidin/alkaline phosphatase conjugate is then added, which will bind to the biotin molecules. Another wash removes excess conjugate. A fluorigenic substrate (4-methylumbelliferyl phosphate) is added, which will be hydrolyzed and the intensity of the emitted fluorescence will allow quantification of the IgG antibody levels to the SARS-CoV-2 RBD protein in the samples. Titers are given in arbitrary units per milliliter (AU/mL) with a cut-off value of 1.95 AU/mL.

#### sVNT assay

Antibody neutralization properties were determined using an “in-house” surrogate virus neutralization test (sVNT) measuring RBD-ACE2 (angiotensin-converting enzyme 2) inhibition (CIGB, Cuba) and confirmed by a live viral neutralization test (Civilian Defense Scientific Research Center, Cuba). In the surrogate neutralization test, vaccinee samples were pre-incubated with peroxidase-conjugated recombinant RBD protein at different dilutions and subsequently the mixture is incubated with ACE-2 protein fixed in the solid phase. Results are given in inhibition percentage. The assay threshold for positivity is 20%. The live neutralization assay is based on the standard virus microneutralization assay. Vero E6 cells are incubated in absence or presence of diluted sera and overlaid with viral suspension. At 96 h post-infection, the cytopathogenic effect (CPE) was evaluated by optical microscopy and the cells stained with neutral red. After three washes neutral red was dissolved in lysis solution and optical density (OD) was measured at 540 nm. The viral neutralizing titer (VNT50) is calculated as the highest serum dilution at which 50 % of the cells remain intact, compared to neutral red incorporation in the control wells (no virus added).

Costar 3591 plates were coated with hFc-ACE2 protein (supplied by the Center for Molecular Immunology, Cuba) at 250ng per well in phosphate-buffered saline (PBS) pH 7.4 for three hours at 37°C. After a wash step with 0.1% (v/v) of Tween-20 in distilled water, the plates were blocked with 2% (w/v) nonfat dry milk powder in PBS for 1h at 37°C. During the coating incubation period, the samples and assay controls were pre-incubated with hFc-RBD-HRP (CIGB, Cuba) conjugate diluted 1:100 000 with 0.2% (w/v) skim milk in PBS, for 1h at 37 °C. A Mab, CBSSRBD-S.8, (CIGBSS, Cuba) with significant neutralization activity against SARS-CoV-2 was used as positive control in this surrogate assay. Human AB Serum (Sigma, USA) was used as a negative control. After the pre-incubation period, 50µL of sample-conjugate mixture is added to the ACE-2 blocked plate and incubated for 1h at 37°C. Unbound reactants are removed by four washes with 0.1% (v/v) of Tween-20 in distilled water. After washing, 50 µL of 3,3’,5,5’-tetramethylbenzidine at 10µg/mL dissolved in phosphate-citrate buffer (0.2M phosphate, 0.1M citrate, pH 5.0) and 0.006% (v/v) hydrogen peroxide were added per well, followed by incubation at room temperature for 10 min. The reaction was stopped by adding Stop solution (2M H_2_SO_4_). Microtiter plates were read at a 450 nm wavelength.

Inhibition values were determined as follows:

Inhibition (%) = (1 - sample optical density value/negative control optical density value) × 100.

Results are given in inhibition percentage. The assay threshold for positivity is 20%. Inhibition values were plotted in GraphPad Prism 8.0.2 and the neutralization titer was defined as the concentration that showed 50% inhibition (inhibitory concentration, IC50) as determined by log (inhibitor) vs. normalized response – variable slope model.

#### VNT50 assay

Neutralization antibody titers were detected by a standard virus microneutralization assay (MN50) using SARS-CoV-2 (CUT2010-2025/Cuba/2020 strain) [28]. Vero E6 cells (2×10^4^ per well) were seeded in 96-well plates one night before use. Human sera were inactivated at 56 °C for 30 min. The serum samples were prepared by two-fold serial dilutions in Eagle Minimal Essential Medium (MEM, Gibco, UK) containing 2 % (v/v) fetal bovine serum (Capricorn, Germany). SARS-CoV-2 strain at 100 TCID50 was incubated in absence or presence of diluted sera for 1 h at 37 °C. Afterwards, Vero E6 cell were overlaid with virus suspension. At 96 h post-infection, the cells were inspected for signs of cytopathogenic effects (CPE) by optical microscopy and stained with neutral red (Sigma, USA). After three washes neutral red was dissolved in lysis solution (50 % ethanol, 1 % acetic acid) for 15 min at 25 °C, and optical density (OD) was detected at 540 nm. The highest serum dilution showing an OD value larger than the cut-off was considered the neutralization titer. The cut-off value is calculated as the average of the OD of the cell control wells divided by two. Viral neutralizing titers (VNT50) were calculated as the highest serum dilution at which 50 % of the cells remain intact according to neutral red incorporation in the control wells (no virus added).

### Statistical analysis

All statistical analyses were performed by GraphPad Prism v. 8.0. In line with common conventions of descriptive statistics, standard deviation (SD) was calculated for mean values and interquartile range (IQR) for median values, geometric mean titer (GMT) and 95% confidence intervals (CIs) were also estimated. We used Wilcoxon matched-pairs signed rank test to compare the pre and post-immunization titers of serum samples and Spearman non-parametric correlation coefficient (Spearman’s rho, r) test to perform inferential analysis among tests results with the categories: none or very weak r < 0.3, weak 0.3 < r <0.5, moderate 0.5 < r < 0.7 and strong r > 0.7. Cohen’s kappa (κ) [9] was calculated to assess the agreement between the test assays with the categories of poor (below 0.00), slight (0.00–0.20), fair (0.21–0.40), moderate (0.41–0.60), substantial (0.61–0.80) and almost perfect (0.81–1.00).

## Results

Antibody titers obtained with the Elecsys^®^ Anti-SARS-CoV-2 S test in Abdala vaccinees are shown in Figure 1. Prior to immunization, only one sample showed detectable anti-RBD antibodies (≥ 0.80 U/mL) by the Roche test. Post-vaccinated sera showed a GMT of 1382 U/mL (95% CI: 850.7-2247), that represents a 3191.7-fold increase compared to the baseline value of 0.43 U/mL obtained before immunization. In Abdala vaccinated individuals, the seroconversion rate 15 days after the last dose was 100% by the Roche test.

**Figure 1:**
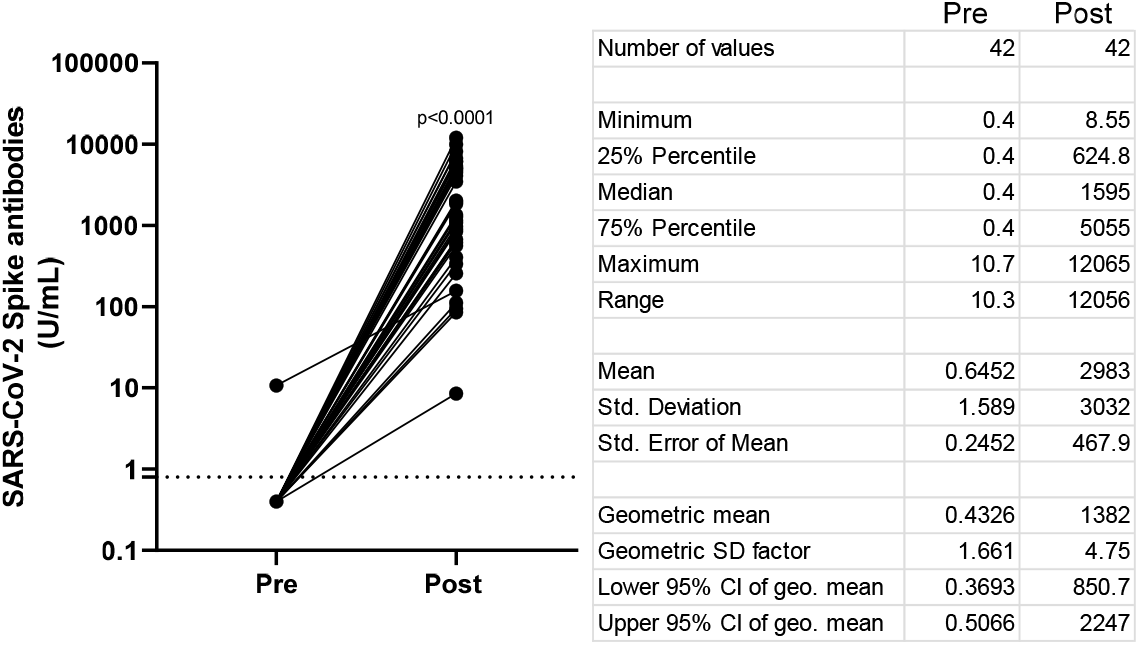
Before-after graph showing SARS-CoV-2 Spike antibody titers obtained by Elecsys® Anti-SARS-CoV-2 S in Abdala vaccinees. Statistical differences between Pre and Post immunized serum groups (P < 0.0001) were determined by Wilcoxon matched-pairs signed rank test. The dotted line represents the cut-off of the test (≥ 0.8 U/mL). The table shows the descriptive statistics of these results.

The degree of agreement between both tests was analyzed in 126 evaluated samples. Results are shown in Table 1. The concordance value (κ) obtained was 0.873 (95% CI, 0.781 to 0.964).

**Table 1.**
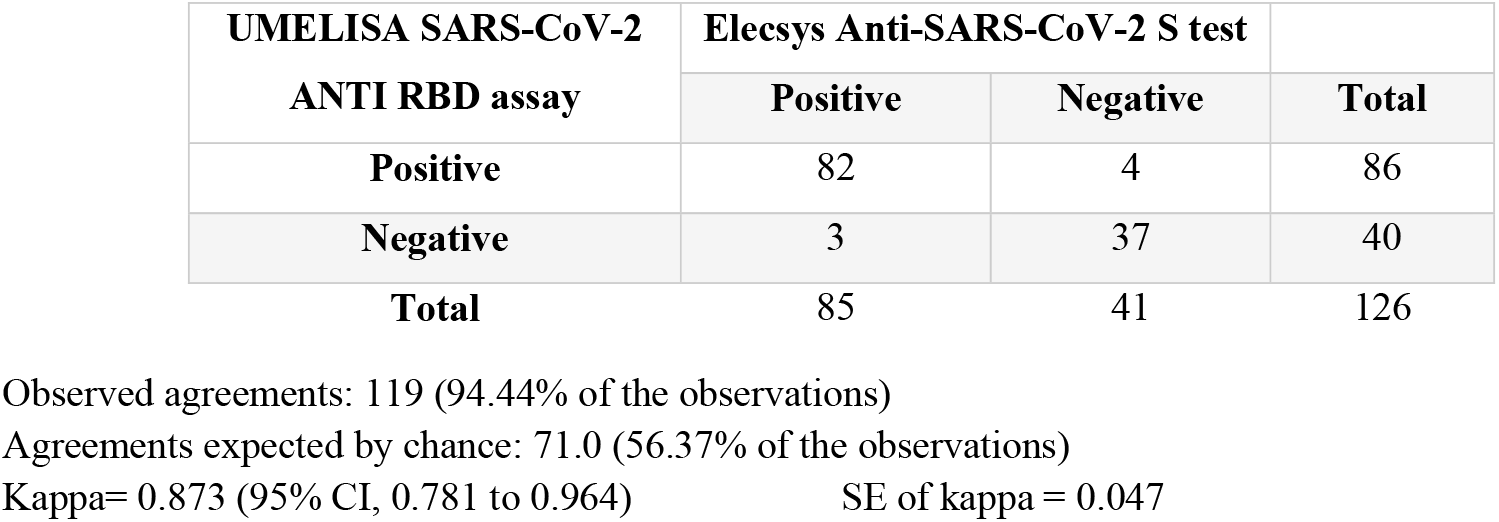
Two-way contingency table for anti-RBD antibody detection using UMELISA SARS-CoV-2 ANTI RBD and Elecsys^®^ Anti-SARS-CoV-2 S (considered the gold standard test).

| UMELISA SARS-CoV-2<br>ANTI RBD assay | Elecsys Anti-SARS-CoV-2 S test |  | Total |
| --- | --- | --- | --- |
|  | Positive | Negative |  |
| Positive | 82 | 4 | 86 |
| Negative | 3 | 37 | 40 |
| Total | 85 | 41 | 126 |

Observed agreements: 119 (94.44% of the observations)
Agreements expected by chance: 71.0 (56.37% of the observations)
Kappa= 0.873 (95% CI, 0.781 to 0.964)
SE of kappa = 0.047

Correlation between the Roche and UMELISA tests is shown in Figure 2. A positive value of r= 0.9589, (CI: 0.9415-0.9711), suggests a strong correlation between both assays.

**Figure 2:**
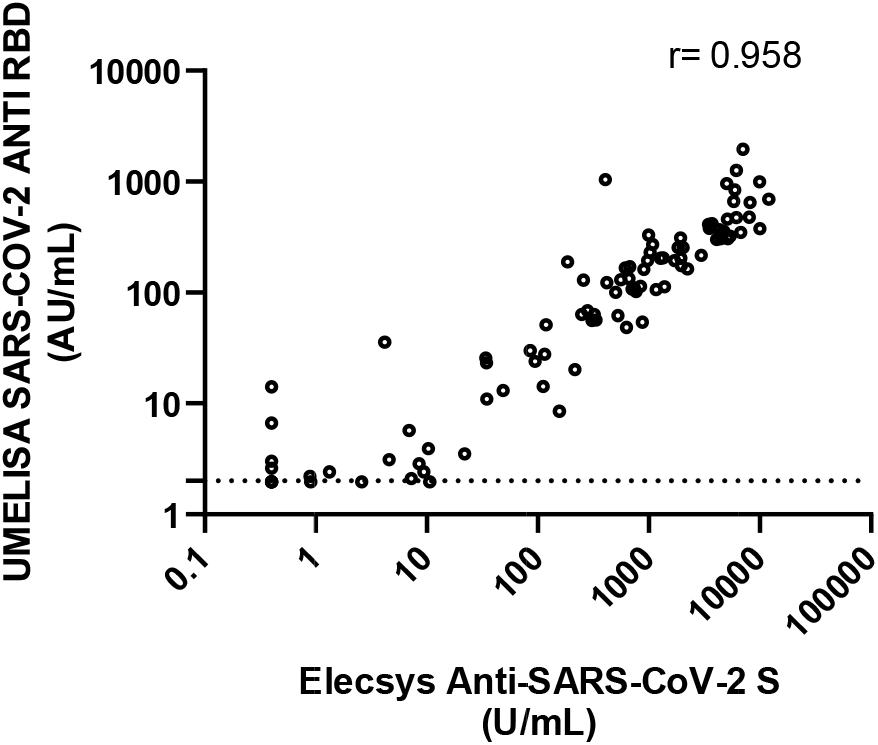
Correlation analysis between antibody titer results obtained by Elecsys^®^ Anti-SARS-CoV-2 S and UMELISA SARS-CoV-2 ANTI RBD in vaccinated individuals. The dotted line represents the test cut-off value (1.95 AU/mL).

Although 10 positive samples by Elecsys^®^ Anti-SARS-CoV-2 S showed inhibition values below 20% by the sVNT assay, a rho value of 0.9111 (95% CI: 0.8748 to 0.9373) was estimated, suggesting a strong positive correlation between both assays (Figure 3).

**Figure 3:**
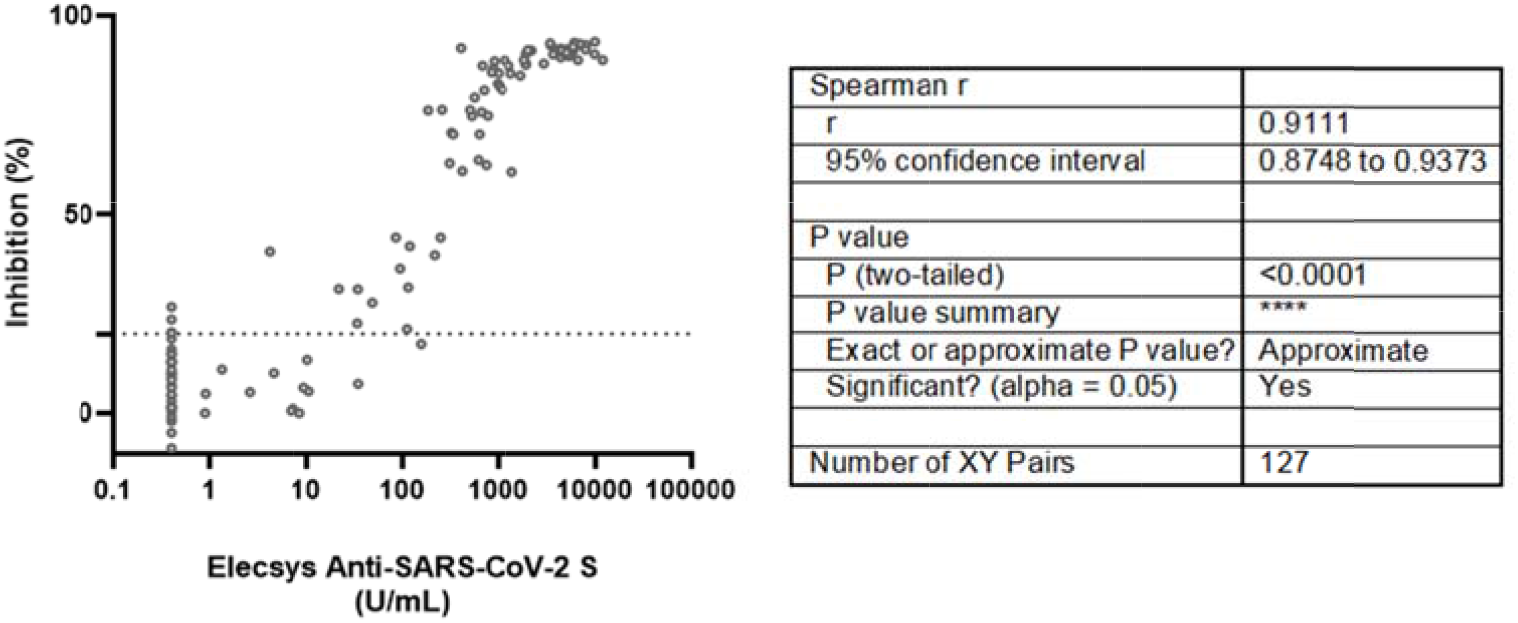
Correlation analysis between antibody titer results obtained by Elecsys® Anti-SARS-CoV-2 S and sVNT assay (% of inhibition) in vaccinated individuals. The dotted line represents the test cut-off value (20%).

The live viral micro-neutralization test also showed a strong positive correlation with the Roche assay (Figure 4). Although five anti-RBD positive samples by the Roche test showed no neutralization titers, there is a positive strong correlation between both tests (r =0.8773).

**Figure 4:**
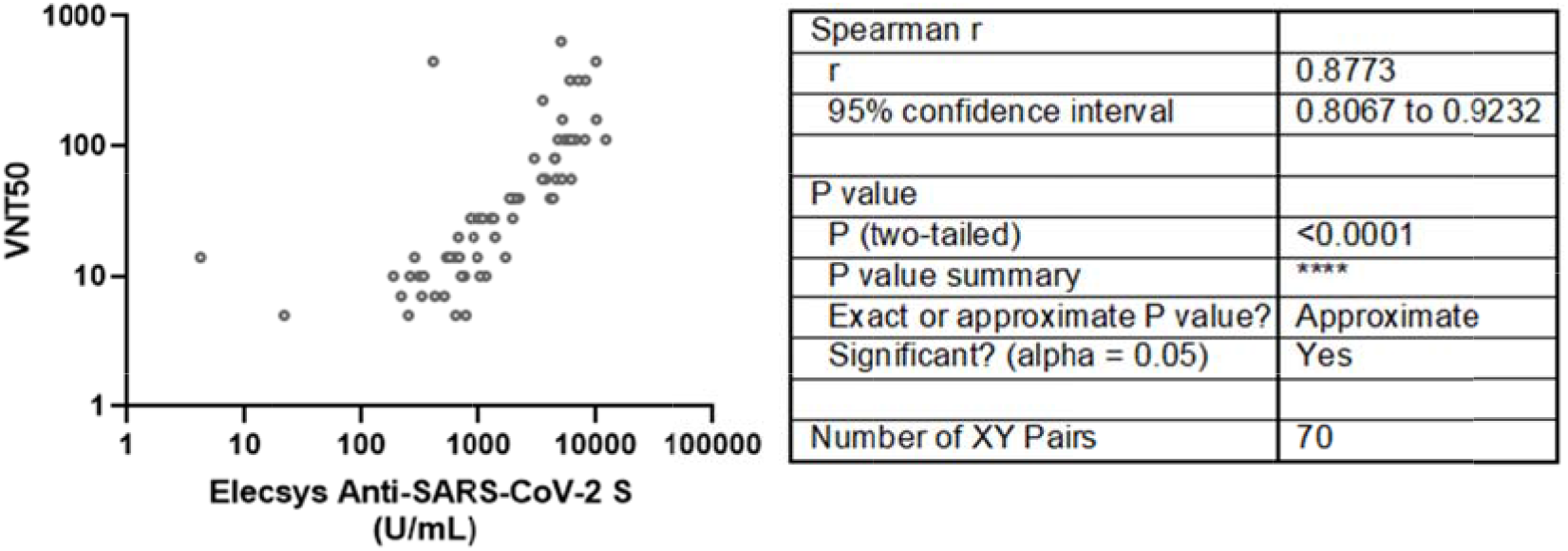
Correlation analysis between antibody titer results obtained by Elecsys® Anti-SARS-CoV-2 S and VNT assay (neutralization titer) in vaccinated individuals.

## Discussion

Since the beginning of the COVID-19 pandemic, many efforts have been displayed to obtain highly effective vaccines capable of eliciting protective immune response, which is essential for the prevention and mitigation of the morbidity and mortality caused by the virus. Therefore vaccines are expected to be decisive in the strategies for stopping, controlling, or slowing the pandemic, fighting the virus, and limiting worldwide transmission of SARS-CoV-2.

Currently, there are several commercially available vaccines, based on different approaches and varying efficacies, such as the mRNA-based BioNtech-Pfizer and Moderna (95% and 94% respectively) [29,30], using an adenovirus vector as AstraZeneca (70% efficacy) [31] and Sputnik V (91.6% efficacy) [32] and inactivated virus as CoronaVac (50% efficacy) [33]. With several billion doses of COVID-19 vaccines already administered [34]; we are witnessing the largest global vaccine deployment in history.

As this study shows, Abdala vaccinees exhibited high antibody titers by the Roche test (antibody titer median of 1595 U/mL). This result was not a surprise, since it has been recognized that mannosylated proteins secreted in *Pichia pastoris* as a host, have the potential to function as adjuvants, favoring in this case an RBD subunit vaccine. In addition there is enhanced antigen presentation and T-cell activation properties upon interaction with receptors in antigen-presenting cells [35].

The antibody titers obtained in Abdala vaccinees were similar to antibody titers previously reported for a commercial vaccine: BNT162b2 from Pfizer-BioNTech using the same test [36].

During serological studies of the Cuban COVID-19 vaccine candidates, throughout the preclinical and clinical phases, the abovementioned Cuban commercial and in-house tests have been used. It is important to note that the proteins used for IgG quantification or the inhibition assays were obtained in mammalian cells, a heterologous expression system with respect to the one used for obtaining the vaccine antigen.

Although the antibody quantification assays used in this study have different principles, and moreover, the Roche sandwich test detects total high affinity antibodies and UMELISA detects IgG type antibodies, both had an almost exact agreement (κ= 0.873) and a strong positive correlation (r=9589).

sVNT and VNT50 assays also showed a positive and strong correlation with the Elecsys® Anti-SARS-CoV-2 S test (rho value of 0.911 and 0.8773 respectively).

## Conclusions

The different tests used (the in-house sVNT and the UMELISA SARS-CoV-2 ANTI RBD assays (Immunoassay Center, Cuba) showed strong positive correlation with the Elecsys® Anti-SARS-CoV-2 S test (Roche, USA) for antibody detection. Since the last is considered the WHO reference standard, the conclusion is that both the in-house and the Cuban UMELISA system are equally accurate for detecting anti-SARS-CoV-2 antibodies. The VNT using live virus also strongly correlates with them, but must be carried out under more stringent biosafety conditions.

Indirectly, the results also show the high immune stimulating capability of the Abdala vaccine, whose vaccinees reach antibody titers by the Elecsys® Anti-SARS-CoV-2 S test similar to those of other approved vaccines like Pfizer-BioNTech when evaluated with the same commercial test.

## Data Availability

All data produced in the present work are contained in the manuscript

## Acknowledgements

The authors would like to express their gratitude to Dr. Loida Torres from the International Health Center “La Pradera” for enabling the use of its facilities and the equipment necessary for the evaluation of the samples.

Research reported here was supported with funds from the BioCubaFarma Enterprise and the Center for Genetic Engineering and Biotechnology in Cuba.

